# Temporal sequence of amyloid and tau PET positivity: *APOE*-ε4 and sex effects, and implications for Alzheimer’s disease progression

**DOI:** 10.1101/2025.07.27.25332202

**Authors:** Marta Milà-Alomà, Isabella Hausle, Kellen K. Petersen, Pamela Thropp, Suzanne E. Schindler, Duygu Tosun, Alzheimer’s Disease Neuroimaging Initiative

**Author notes:** Correspondence to: Duygu Tosun, PhD, Full address: Department of Radiology and Biomedical Imaging, University of California San Francisco, San Francisco, CA, USA. Alzheimer’s Disease Neuroimaging Initiative (ADNI): Data used in preparation of this article were obtained from the Alzheimer’s Disease Neuroimaging Initiative (ADNI) database (adni.loni.usc.edu). As such, the investigators within the ADNI contributed to the design and implementation of ADNI and/or provided data but did not participate in analysis or writing of this report. A complete listing of ADNI investigators can be found at:http://adni.loni.usc.edu/wpcontent/uploads/how_to_apply/ADNI_Acknowledgement_List.pdf.

## Abstract

**Importance:** Alzheimer’s disease (AD) progression varies widely among individuals. Identifying factors influencing timing of pathology and clinical progression is crucial for optimizing early intervention trials.

**Objective:** To investigate how estimated age at amyloid and tau PET positivity, and the time interval between amyloid and tau PET positivity (“amyloid-tau interval”), relate to symptom onset and clinical progression, and whether *APOE-*ε4 status and sex modify these associations.

**Design:** Longitudinal observational study.

**Setting:** Multicenter study using data from the Alzheimer’s Disease Neuroimaging Initiative (ADNI).

**Participants:** ADNI participants with at least one positive amyloid PET scan (n=792) or at least one positive tau PET scan (n=212) were included. Both cognitively unimpaired and impaired individuals were included and all had information on sex, *APOE-*ε4 status, and longitudinal cognitive assessments.

**Exposures:** ^18^F-florbetapir or ^18^F-florbetaben amyloid PET, ^18^F-flortaucipir tau PET, CDR global, CDR-SB.

**Main outcomes and measures:** We examined the influence of *APOE*-ε4 status, sex, and their interaction on the age at biomarker positivity and the amyloid–tau interval. Accelerated Failure Time (AFT) models were used to predict time to symptom onset based on biomarker positivity age and the amyloid–tau interval. Linear mixed-effects (LME) models evaluated differences in the rate of cognitive decline over five years following symptom onset, by biomarker positivity age and amyloid–tau interval duration. Additional models included interaction terms with sex or *APOE*-ε4 status.

**Results:** *APOE-*ε,4 carriers and women had earlier amyloid and tau PET positivity ages. *APOE-*ε,4 carriers, women, and those with older age at amyloid PET positivity had a shorter amyloid-tau interval. An older age at amyloid or tau PET positivity and a shorter amyloid-tau interval predicted earlier symptom onset. An older age at amyloid or tau PET positivity was also associated with slower rate of cognitive decline after symptom onset. The amyloid-tau interval did not influence time until symptom onset after tau PET positivity or rate of cognitive decline.

**Conclusions and relevance:** *APOE-*ε4 and sex influenced the timing of amyloid and tau PET positivity and the amyloid-tau interval, which in turn affected symptom onset and clinical progression. These factors should guide the identification of amyloid-positive individuals at highest risk of rapid AD progression, enabling more efficient selection of participants for clinical trials.

## Introduction

Alzheimer’s disease (AD) is characterized by the accumulation of amyloid-β (Aβ) plaques and tau neurofibrillary tangles, which begin years before development of clinical symptoms^1,2^. Amyloid deposition typically precedes widespread tau aggregation^1^, and both pathologies accumulate relatively consistently across individuals^1,3,4^; however, interindividual variability exists in the onset and temporal sequence of amyloid and tau pathologies and their contribution to clinical progression^3,4^. Understanding the factors driving this variability is essential for improving disease staging, prognosis, and treatment timing.

Positron emission tomography (PET) imaging enables the *in vivo* quantification of amyloid and tau pathologies. PET-based modeling can estimate an individual’s age at which a specific biomarker positivity threshold is crossed^3,4^. These estimated ages for PET positivity offer a valuable tool for studying the temporal dynamics of AD pathophysiology at an individual level and identifying factors influencing key pathological milestones. Anchoring analyses to an individual’s biological disease timeline may also enhance the prediction of clinical outcomes, such as symptom onset or rate of cognitive decline.

Although advanced age is the primary risk factor for AD, genetic and demographic factors such as the apolipoprotein E (*APOE*) genotype and sex significantly contribute to disease risk and progression^5^. The *APOE-* ε4 allele, the strongest genetic risk factor for late-onset AD, is consistently associated with earlier amyloid and tau accumulation, and earlier development of clinical symptoms^6^. Sex differences have also been reported in AD prevalence and progression, with men showing less vulnerability to tau pathology and slower clinical decline than women^7–12^. Furthermore, interactions between *APOE*-ε,4 and sex have been observed to influence AD risk and progression^13–15^.

Despite these established connections, it remains unclear to what extent *APOE-*ε4 status and sex, independently and interactively, influence the timing of amyloid and tau PET positivity, and how these temporal dynamics influence the disease course. While amyloid positivity is necessary, the emergence of tau and subsequent cognitive decline signal imminent clinical progression^16–20^. A better understanding of factors influencing the time interval from reaching amyloid positivity to subsequent tau positivity, which we term “the amyloid-tau interval,” could identify individuals at higher risk of rapid progression once amyloid positivity is established. This has significant implications for early-stage clinical trials, which often enroll participants based solely on amyloid burden. Refining participant selection by considering both amyloid burden and individual trajectories toward tau positivity and symptom onset could improve trial efficiency and therapeutic impact. Investigating how *APOE* genotype and sex impact these key events can offer valuable insight into the mechanisms driving variability in tau emergence, with direct implications for prognostic modeling and selection of individuals for early intervention in clinical trials.

This study investigated how estimated ages at amyloid and tau PET positivity, and the amyloid-tau interval, relate to symptom onset and clinical progression. We also assessed the independent and interactive effects of *APOE-*ε4 status and sex on these associations. By characterizing how these factors modulate the temporal sequence of events in AD pathogenesis, we aim to better understand individual variability in AD progression and its potential clinical consequences.

## Methods

### 1. Study participants

This study included cognitively unimpaired (CU) and cognitively impaired (CI) participants from the Alzheimer’s Disease Neuroimaging Initiative (ADNI) study (adni.loni.usc.edu). ADNI is a public-private partnership focused on identifying biomarkers for mild cognitive impairment (MCI) and early AD progression^21,22^. All participants provided written consent.

For this study, we required at least one positive amyloid PET scan or tau PET scan to estimate biomarker positivity ages (**eMethods in Supplement 1**). All included participants had longitudinal cognitive assessments, self-identified sex, and *APOE* genotype data, which was part of the standard ADNI protocol.

### 2. Amyloid and tau PET measures

Amyloid PET imaging used ^18^F-florbetapir (FBP) or ^18^F-florbetaben (FBB) with standardized ADNI protocols^23^. Global cortical standardized uptake value ratio (SUVR) was calculated by averaging PET uptake across cortical regions (frontal, cingulate, parietal, and lateral temporal) and normalizing to a composite reference region (whole cerebellum, brainstem/pons, and subcortical eroded white matter)^24^. Amyloid positivity was defined as SUVR>0.78 for FBP or SUVR>0.74 for FBB, per ADNI guidelines (ADNI_UCBerkeley_AmyloidPET_Methods_v2_2023-06-29.pdf).

Tau PET imaging used ^18^F-flortaucipir (FTP) with standardized ADNI protocols. Mesial-temporal meta-region of interest (ROI) uptake (including entorhinal, parahippocampus, and amygdala) was normalized to the inferior cerebellar grey matter uptake^25^. We determined mesial-temporal tau PET positivity threshold of 1.41 SUVR using a two-component Gaussian mixture model (GMM) applied to cross-sectional mesial-temporal FTP-PET SUVR from the entire ADNI study (*n*=907).

### 3. Clinical and neuropsychological assessments

Participants underwent comprehensive clinical assessments, including collateral interviews, neurological examinations, and the Clinical Dementia Rating® (CDR®) and the CDR-Sum of Boxes (CDR-SB)^26^. Individuals were classified as CI if they had two consecutive visits with a CDR>0 and a CDR>0 in their last visit. Participants who reverted to a CDR=0 were excluded. Symptom onset was defined as the first visit with a CDR>0.

### 4. Statistical analyses

We estimated ages at amyloid and tau PET positivity using a previously validated method for FBP PET and FTP PET in ADNI^3,27^. We extended this method to FBB PET (in 19.9% of participants with amyloid PET), showing similar performance (**eMethods and eFigure 1 in Supplement 1**).

We used linear regression to assess the independent effects of *APOE-*ε4 status (*APOE-*ε4 carrier *vs. APOE-* ε4 non-carrier) and sex on estimated amyloid and tau PET positivity ages. We then examined the association between these biomarker positivity ages, adjusting for sex and *APOE-*ε4 status, and included the *APOE-*ε4 status by sex interaction term in additional models.

The amyloid-tau interval was calculated by subtracting the estimated age at amyloid PET positivity from the estimated age at tau PET positivity. We used linear regression to examine the effects of *APOE-*ε4 status, sex, and their interaction on the amyloid-tau interval. Additional models included the estimated age at amyloid PET positivity and its interaction with *APOE-*ε4 status, sex, and the three-way interaction.

We used Accelerated Failure Time (AFT) survival models with a Weibull distribution to assess how biomarker positivity ages and the amyloid–tau interval influenced time until symptom onset (from CDR=0 to CDR>0) following amyloid or tau PET positivity. The time-to-event variable was the difference between age at CDR>0 (or the last cognitive assessment for non-converters) and estimated amyloid or tau PET positivity age. Sex, *APOE-*ε4 status, and educational attainment were included as covariates. Additional models tested interactions between biomarker positivity ages or amyloid–tau interval with sex, *APOE-*ε4 status, and their three-way interaction.

Lastly, we used linear mixed effect (LME) models with random slopes and intercepts to assess the effect of biomarker positivity age or amyloid-tau interval on the rates of cognitive decline, measured by CDR-SB, over five years following symptom onset (defined as the first visit with a CDR>0). We grouped participants by biomarker positivity age (<65, 65-75, and >75 years). We also grouped participants by the amyloid-tau interval: “concurrent amyloid-tau” (−4 years ≤ interval ≤ 4 years, reflecting cumulative mean absolute error for estimated biomarker positivity ages; see **eMethods in Supplement 1**), “Amyloid first <10 years” (4-10 years), and “Amyloid-first β10 years”. Six participants with tau PET positivity preceding amyloid PET positivity (interval ≤ −4 years) were excluded from modeling but included descriptively. Interaction terms between time since symptom onset and biomarker age group or interval group were evaluated. Next, pairwise contrasts were conducted to compare group-specific cognitive decline rates. We repeated these analyses adding interactions between time, biomarker positivity age or amyloid-tau interval group and sex or *APOE*-ε,4 status.

All statistical analyses and figure generation were performed using R software (R version 4.2.2).

## Results

### 1. Study participants characteristics and estimated ages at amyloid and tau PET positivity

Our study included 792 participants with estimated amyloid PET ages and 212 with estimated tau PET positivity age; 152 participants had both estimates. Demographic and clinical characteristics were similar between these subgroups (**Table 1**).

**Table 1.** Participant characteristics at first PET acquisition.

|  | Cohort with estimated age at amyloid PET positivity (n=792) | Cohort with estimated age at tau PET positivity (n=212) | Cohort with PET positivity age for both amyloid and tau (n=152) |
| --- | --- | --- | --- |
| Age, years | 74.3 ± 7.3 | 76.3 ± 7.8 | 73.9 ± 6.8 |
| Women, n (%) | 399 (50.4) | 103 (48.6) | 74 (48.7) |
| *APOE*-ε4 carriers, n (%) | 493 (62.2) | 127 (59.9) | 104 (68.4) |
| Educational attainment (High school or less / College / Postgraduate), n (%) | 120 (15.2) / 350 (44.2) / 322 (40.7) | 20 (9.4) / 103 (48.6) / 89 (42.0) | 17 (11.2) / 74 (48.7) / 61 (40.1) |
| Race (Black/White/Other), n (%) | 39 (4.9) / 728 (91.9) / 25 (3.2) | 8 (3.8) / 189 (89.2) / 15 (7.1) | 6 (3.9) / 136 (89.5) / 10 (6.6) |
| CDR 0/0.5/>0.5, n (%) | 248 (31.3) / 453 (57.2) / 91 (11.5) | 72 (34.0) / 113 (53.3) / 27 (12.7) | 54 (35.5) / 91 (59.9) / 7 (4.6) |
| CDR-SB | 1.75 ± 2.11 | 1.83 ± 2.25 | 1.25 ± 1.49 |
| Amyloid PET, Centiloid | 67.6 ± 35.5 | 68.3 ± 46.4 | 68.9 ± 30.8 |
| Amyloid PET-positive<sup>a</sup>, n (%) | 696 (87.9) | 158 (79) | 137 (90.1) |
| Mesial-temporal tau PET, SUVR | 1.51 ± 0.44 | 1.63 ± 0.23 | 1.66 ± 0.25 |
| Mesial-temporal tau PET-positive<sup>b</sup>, n (%) | 99 (47.1) | 180 (84.9) | 78 (51.3) |
| Amyloid PET tracer (<sup>18</sup>F-florbetapir/<sup>18</sup>F-florbetaben), n (%) | 634 (80.1) / 158 (19.9) | 125 (62.5) / 75 (37.5) | 89 (58.6) / 63 (41.4) |

Data are presented as mean ± standard deviation or n (%). 210 participants with estimated amyloid PET positivity age had a concurrent tau PET scan, and 200 participants with estimated tau PET positivity age had a concurrent amyloid PET scan. 152 participants had estimates for both amyloid and tau PET positivity age and overlapped between the two cohorts. <sup>a</sup>Amyloid PET positivity threshold: > 0.78 SUVR for FBP scans and > 0.74 SUVR for FBB scans; <sup>b</sup>Mesial-temporal tau PET positivity threshold: >1.41 SUVR.

The mean estimated amyloid and tau PET positivity ages were 63.3±10.2 years and 69.9±9.9 years, respectively. Both *APOE*-ε,4 carriership and female sex were significantly associated with earlier biomarker positivity ages (all *P*<0.05; **Figure 1A and 1B**). A significant *APOE*-ε,4 status by sex interaction was found for amyloid PET positivity age (*P*=0.040), with the association between *APOE*-ε,4 and earlier amyloid PET positivity age being stronger in women (**Figure 1A**). No significant interaction effect was observed for tau PET positivity age (*P*=0.52; **Figure 1B**).

**Figure 1.**
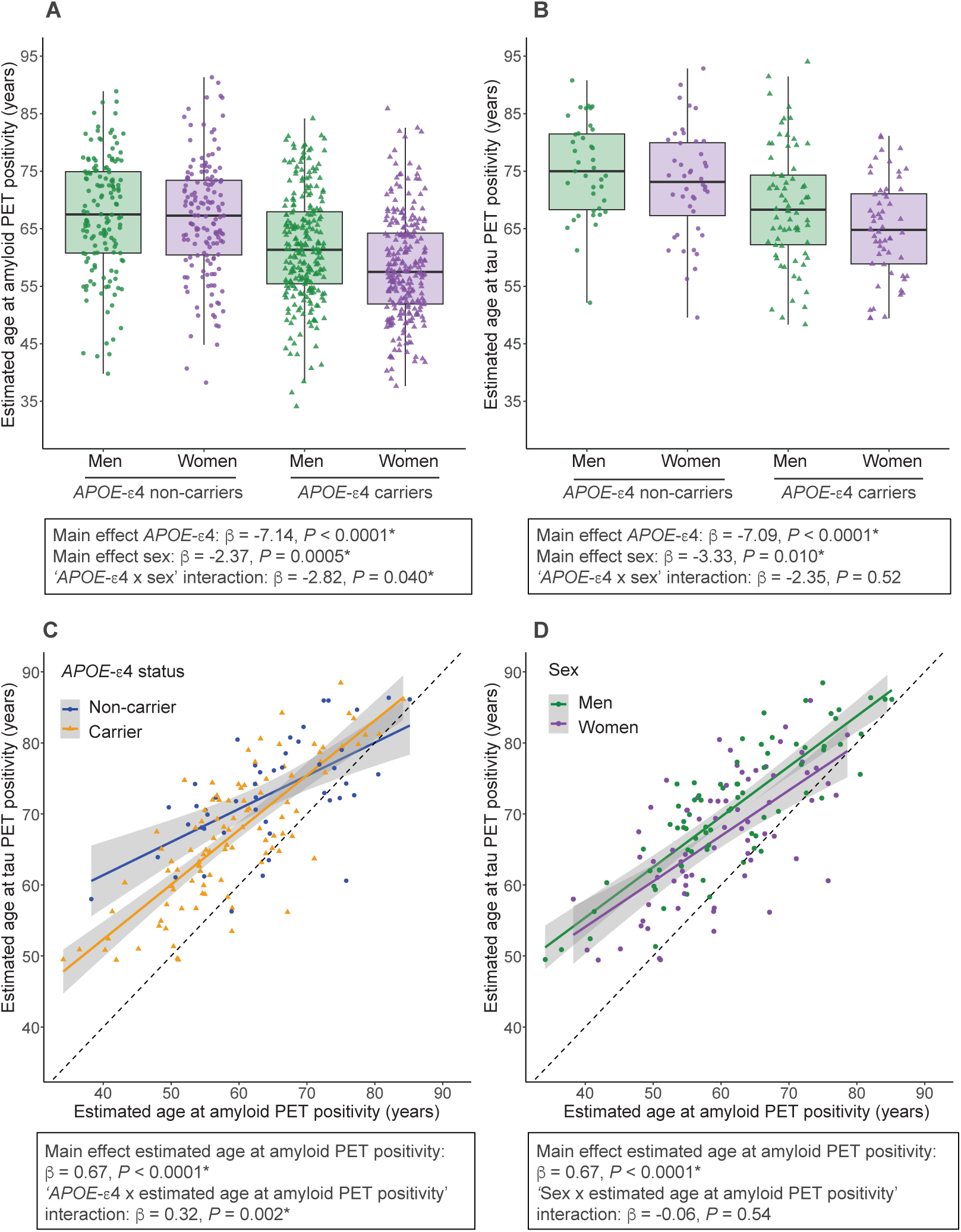
Estimated ages at amyloid and tau PET positivity by *APOE*-ε.4 status and sex A, B. Estimated ages at amyloid (A) or tau (B) PET positivity by *APOE*-ε4 status and sex. Box plots depict median (horizontal bar), IQR (hinges), and 1.5 × IQR (whiskers). C, D. Scatter plots show the association between estimated age at amyloid PET positivity and at tau PET positivity by *APOE*-ε4 status (C) and sex (D). Solid lines indicate the regression line and 95% CIs. The dashed diagonal line represents the identity line (y = x), indicating equal ages at amyloid and tau PET positivity. Beta coefficients and *P*-values from linear regression models are reported for the main effects of *APOE*-ε4 status, sex or estimated age at amyloid PET positivity, and the interaction terms with *APOE*-ε4 and sex. All models included *APOE*-ε4 status and sex as covariates. Interaction terms were added in additional models.

Among 152 participants with both estimates, earlier amyloid PET positivity age was significantly associated with an earlier tau PET positivity age (β=0.67, *P*<0.0001). This association was significantly modified by *APOE*-ε,4 status (*P*=0.002; **Figure 1C**), showing a stronger effect in *APOE*-ε,4 carriers. Sex did not moderate this association (*P*=0.54, **Figure 1D**), nor did the three-way interaction between amyloid PET positivity age, *APOE*-ε,4 status, and sex (*P*=0.92). **eTable 1 in Supplement 1** shows participant characteristics by estimated biomarker positivity age groups (<65, 65-75 or >75 years old).

### 2. Amyloid-tau interval: Distribution and differences by *APOE*-ε,4 status and sex

The mean amyloid-tau interval was 8.2±6.8 years. The amyloid-tau interval distribution (**Figure 2A**) showed 34 individuals (22.4%) had a “concurrent amyloid-tau” interval (−4 years ≤ interval ≤4 years). Most individuals (n=112; 73.7%) showed amyloid PET positivity preceding tau PET positivity (‘amyloid-first’ group), with 65 (58.0%) of them having an interval exceeding 10 years. Six individuals (3.9%) had tau PET positivity before amyloid PET positivity (‘tau-first’ group). Sex and *APOE*-ε,4 status distribution or cognitive status did not differ across amyloid-interval groups **(eTable 2 in Supplement 1).**

**Figure 2.**
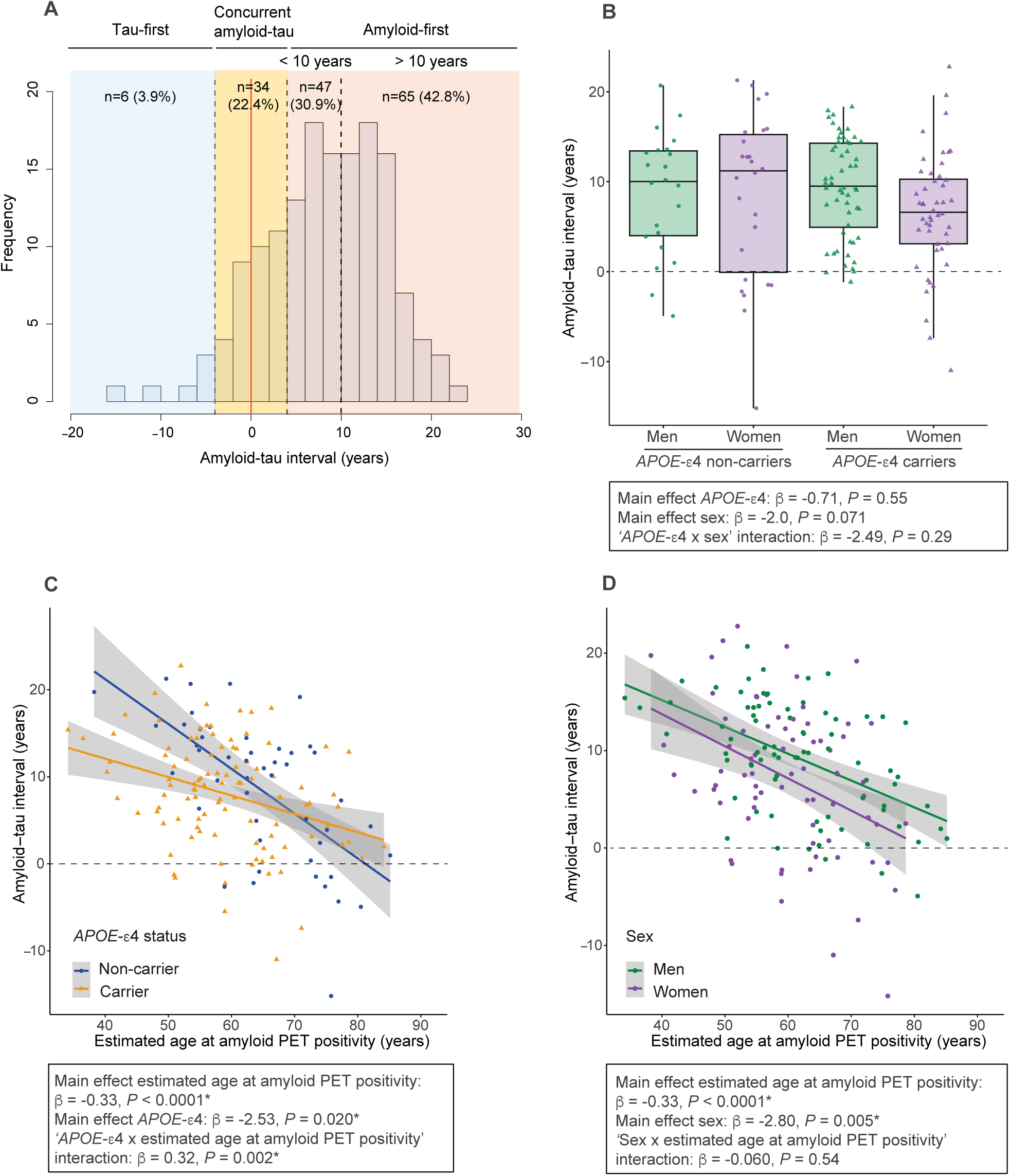
Characteristics of the amyloid-tau interval and effect of *APOE*-ε,4 status and sex A. Distribution of the amyloid-tau interval. The “concurrent amyloid-tau” group was defined by an interval of ± 4 years, reflecting cumulative mean absolute error for estimated positivity ages (see Supplementary Methods). Participants that became amyloid-positive first and tau-positive 4 to 10 years later were classified as “Amyloid first <10 years”, and participants that became amyloid-positive first and tau-positive more than 10 years later were classified as “Amyloid-first ≥10 years”. B. Amyloid-tau interval by *APOE*-ε4 status and sex. Box plots depict median (horizontal bar), IQR (hinges), and 1.5 × IQR (whiskers). C, D-Scatter plots show the association between estimated age at amyloid PET positivity and the amyloid-tau interval by *APOE*-ε4 status (C) and sex (D). Solid lines indicate the regression line and 95% CIs. The dashed horizontal line at 0 years indicates concurrent amyloid and tau PET positivity. Positive values reflect individuals who became tau PET positive after amyloid PET positivity; negative values indicate tau PET positivity preceding amyloid PET positivity. Beta coefficients and *P*-values from linear regression models are reported for the main effects of *APOE*-ε4 status, sex or estimated age at amyloid PET positivity, and the interaction terms with *APOE*-ε4 and sex. All models included *APOE*-ε4 status and sex as covariates. Interaction terms were added in additional models.

*APOE*-ε,4 status, sex, or their interaction did not significantly influence the amyloid-tau interval (**Figure 2B**). However, when amyloid PET positivity age was included in the model, an older age at amyloid PET positivity was significantly associated with a shorter amyloid-tau interval (β= −0.33, *P*<0.0001). In this extended model, *APOE*-ε,4 carriers had a significantly shorter amyloid-tau interval than non-carriers (β=-2.53, *P*=0.020; **Figure 2C**), and women had a shorter interval than men (β=-2.80, *P*=0.005; **Figure 2D**). A significant interaction between *APOE-*ε4 status and estimated age at amyloid PET positivity was observed (*P*=0.002), indicating that the association between older amyloid PET positivity age and shorter amyloid-tau interval was more pronounced in *APOE-*ε4 non-carriers (**Figure 2C**). The interaction between amyloid PET positivity age and sex was not significant (*P*=0.54; **Figure 2D**). The three-way interaction was also not significant (*P*=0.92).

### 3. Effects of biomarker positivity ages and amyloid–tau interval on symptom onset and clinical progression

#### 3.1 Association with time until symptom onset

AFT survival models assessed factors influencing the time until symptom onset (CDR>0) following amyloid or tau PET positivity. In a basic model not including biomarker positivity ages or the amyloid-tau interval (Model 1; **Table 2**), the time until symptom onset was longer for women after amyloid (23% longer; *P*=0.047) and tau (72% longer; *P*=0.003) PET positivity. This sex effect was significantly modified by *APOE-*ε4 status (*P*=0.021; Model 2; **Table 2**), with a 66% longer time until symptom onset after amyloid PET positivity for women *APOE-ε4* carriers compared to men carriers. No significant sex by *APOE-*ε4 interaction was observed after tau PET positivity (*P*=0.50; Model 2; **Table 2**). Neither *APOE-*ε4 status nor educational attainment independently influenced the time until symptom onset.

**Table 2.** Effects of estimated biomarker positivity age and amyloid-tau interval on progression rate.

|  | Time to conversion after amyloid PET positivity |  | Time to conversion after tau PET positivity |  |
| --- | --- | --- | --- | --- |
|  | Time ratio [95% CI] | *P*-value | Time ratio [95%CI] | *P*-value |
| **Effect of estimated biomarker positivity age on time until cognitive impairment** |  |  |  |  |
| **Model 1:** |  |  |  |  |
| *APOE*-ε4 status (carrier) | 1.05 [0.86-1.28] | 0.65 | 1.17 [0.80-1.71] | 0.41 |
| Sex (women) | 1.23 [1.00-1.51] | **0.047\*** | 1.72 [1.21-2.45] | **0.003\*** |
| Educational attainment (college) | 1.04 [0.76-1.42] | 0.82 | 1.07 [0.51-2.26] | 0.86 |
| Educational attainment (postgraduate) | 1.22 [0.87-1.70] | 0.25 | 1.36 [0.66-2.82] | 0.40 |
| **Model 2: Model 1 + 'sex x *APOE*-ε4 status' interaction** |  |  |  |  |
| Sex (women) x *APOE*-ε4 status (carrier) | 1.66 [1.08-2.56] | **0.021\*** | 1.33 [0.59-3.02] | 0.50 |
| **Model 3: Model 1 + age at biomarker positivity** |  |  |  |  |
| Biomarker positivity age | 0.96 [0.95-0.97] | **<0.0001** | 0.97 [0.95-0.99] | **0.002\*** |
| *APOE*-ε4 status (carrier) | 0.87 [0.75-1.02] | 0.085 | 1.08 [0.78-1.50] | 0.63 |
| Sex (women) | 1.03 [0.89-1.21] | 0.68 | 1.29 [0.90-1.84] | 0.17 |
| Educational attainment (college) | 1.02 [0.81-1.28] | 0.87 | 0.77 [0.39-1.53] | 0.46 |
| Educational attainment (postgraduate) | 1.23 [0.96-1.58] | 0.10 | 1.02 [0.54-1.94] | 0.95 |
| **Model 4: Model 3 + 'age at biomarker positivity x *APOE*-ε4 status' interaction** |  |  |  |  |
| Biomarker positivity age x *APOE*-ε4 status (carrier) | 1.00 [0.98-1.01] | 0.81 | 1.00 [0.97-1.04] | 0.97 |
| **Model 5: Model 3 + 'age at biomarker positivity x sex' interaction** |  |  |  |  |
| Biomarker positivity age x sex | 1.00 [0.98-1.01] | 0.60 | 0.99 [0.95-1.03] | 0.48 |
| **Model 6: Model 3 + 'age at biomarker positivity x *APOE*-ε4 status x sex' interaction** |  |  |  |  |
| Biomarker positivity age x sex x *APOE*-ε4 status | 0.97 [0.93-1.00] | 0.081 | 1.04 [0.95-1.13] | 0.42 |
| **Effect of amyloid-tau interval on time until cognitive impairment** |  |  |  |  |
| **Model 1** |  |  |  |  |
| Amyloid-tau interval | 1.06 [1.0-1.08] | **<0.0001** | 0.98 [0.96-1.0] | 0.11 |
| Sex (women) | 1.34 [1.0-1.72] | **0.024\*** | 1.43 [0.94-2.18] | 0.093 |
| *APOE*-ε4 status (carrier) | 1.14 [0.9-1.45] | 0.27 | 1.11 [0.74-1.66] | 0.62 |
| Educational attainment (college) | 0.95 [0.6-1.50] | 0.83 | 1.12 [0.54-2.31] | 0.76 |
| Educational attainment (postgraduate) | 1.03 [0.6-1.63] | 0.89 | 1.14 [0.55-2.39] | 0.73 |
| **Model 2: Model 1 + amyloid-tau interval x sex interaction** |  |  |  |  |
| Amyloid-tau interval x sex (women) | 0.95 [0.90-0.99] | **0.010\*** | 0.94 [0.89-0.99] | **0.028\*** |
| **Model 3: Model 1 + 'amyloid-tau interval x *APOE*-ε4 status' interaction** |  |  |  |  |
| Amyloid-tau interval x *APOE*-ε4 status interaction | 0.98 [0.90-1.01] | 0.14 | 0.98 [0.94-1.03] | 0.40 |
| **Model 4: Model 1 + biomarker positivity age** |  |  |  |  |
| Amyloid-tau interval | 1.05 [1.03-1.06] | **<0.0001** | 0.99 [0.97-1.02] | 0.54 |
| Biomarker positivity age | 0.98 [0.96-0.99] | **0.003\*** | 0.97 [0.94-0.99] | **0.013\*** |
| Sex (women) | 1.17 [0.92-1.50] | 0.21 | 1.14 [0.75-1.74] | 0.53 |
| *APOE*-ε4 status (carrier) | 1.08 [0.86-1.35] | 0.52 | 1.00 [0.69-1.46] | 0.10 |
| Educational attainment (college) | 0.77 [0.49-1.20] | 0.24 | 0.82 [0.40-1.70] | 0.60 |
| Educational attainment (postgraduate) | 0.91 [0.60-1.38] | 0.65 | 0.97 [0.49-1.92] | 0.93 |
| **Model 5: Model 4 + 'amyloid-tau interval x sex' interaction** |  |  |  |  |
| Amyloid-tau interval x sex (women) | 0.97 [0.94-1.01] | 0.15 | 0.97 [0.92-1.03] | 0.29 |
| **Model 6: Model 4 + 'amyloid-tau interval x *APOE*-ε4 status' interaction** |  |  |  |  |
| 'Amyloid-tau interval x *APOE*-ε4 status' interaction | 0.98 [0.95-1.01] | 0.14 | 0.98 [0.94-1.02] | 0.26 |

Accelerated Failure Time (AFT) models were used to examine the main effects of the estimated biomarker positivity age or the amyloid-tau interval along with sex, *APOE*-ε4 status and educational attainment, on the time from amyloid or tau PET positivity to conversion from CDR = 0 to CDR > 0. Interaction terms with sex or *APOE*-ε4 were added in separate models. All models included sex, *APOE*-ε4 status and educational attainment as covariates. A time ratio > 1 indicates slower progression (longer time to impairment), whereas a time ratio < 1 indicates faster progression (shorter time to impairment).

Older age at biomarker positivity was associated with a shorter time until symptom onset; as the age at amyloid or tau PET positivity increased, each year of age corresponded to a 4% (*P*<0.0001) or 3% (*P*=0.002) shorter time until symptom onset, respectively (Model 3; **Table 2 and eFigure 3A and 3B in Supplement 1**). These effects were not significantly modified by sex, *APOE*-ε4 status, or their three-way interaction (Models 4-6; **Table 2**). Notably, once biomarker positivity age was included in the models (i.e., Model 1 vs Model 3), the independent effect of sex was no longer significant, suggesting that the observed longer time to symptom onset in women was largely accounted for by their earlier biomarker positivity age.

Next, we examined the effect of the amyloid-tau interval on the time until symptom onset. A shorter amyloid-tau interval was associated with 6% shorter time until symptom onset for each year of age following amyloid PET positivity (*P*<0.0001), independent of the amyloid PET positivity age (Models 1 and 4; **Table 2 and eFigure 3C in Supplement 1**). Conversely, the amyloid-tau interval had no significant effect on the time to symptom onset after tau PET positivity (*P*=0.11; Models 1 and 4; **Table 2 and eFigure 3D in Supplement 1**). Older tau PET positivity age remained a significant predictor of shorter time to symptom onset (*P*=0.013), irrespective of the amyloid-tau interval (Model 4; **Table 2**). While significant interactions with sex were initially observed, suggesting attenuation of the protective effect of a longer interval in women, these interactions became non-significant when amyloid or tau PET positivity age were included (Models 2 and 5; **Table 2**). *APOE-*ε4 status did not significantly modify these associations (Models 3 and 6; **Table 2**). Three-way interactions could not be evaluated due to sample size limitations.

#### 3.2 Association with clinical progression

Next, we assessed differences in the rates of cognitive decline after symptom onset, defined as the first visit with a CDR>0, across biomarker positivity ages or amyloid-tau interval groups of CI individuals (see **eTable 3 in Supplement 1** for participant characteristics).

There were significant differences across amyloid and tau PET positivity age groups in the rates of cognitive decline after symptom onset (time by amyloid age group interaction *P*=0.005 and time by tau age group interaction *P*=0.026; **Figure 3A and 3B and eTable 4 in Supplement 1**). Post-hoc pairwise comparisons showed that, after symptom onset, individuals who became amyloid PET positive before age 65 declined significantly faster than those >75 years (slope: 0.96 [0.84–1.09] vs. 0.41 [0.08–0.73]; *P*=0.005). A similar pattern was observed when grouping individuals by tau PET positivity age (slope: 1.07 [0.74–1.39] for < 65 vs. 0.39 [0.03–0.76] for >75; *P*=0.020; **Figure 3A and 3B and eTable 4 in Supplement 1**). In contrast, there was no significant effect of the amyloid-tau interval on the rates of cognitive decline after symptom onset (time by interval group interaction *P*=0.41; **Figure 3C and eTable 4 in Supplement 1**).

**Figure 3.**
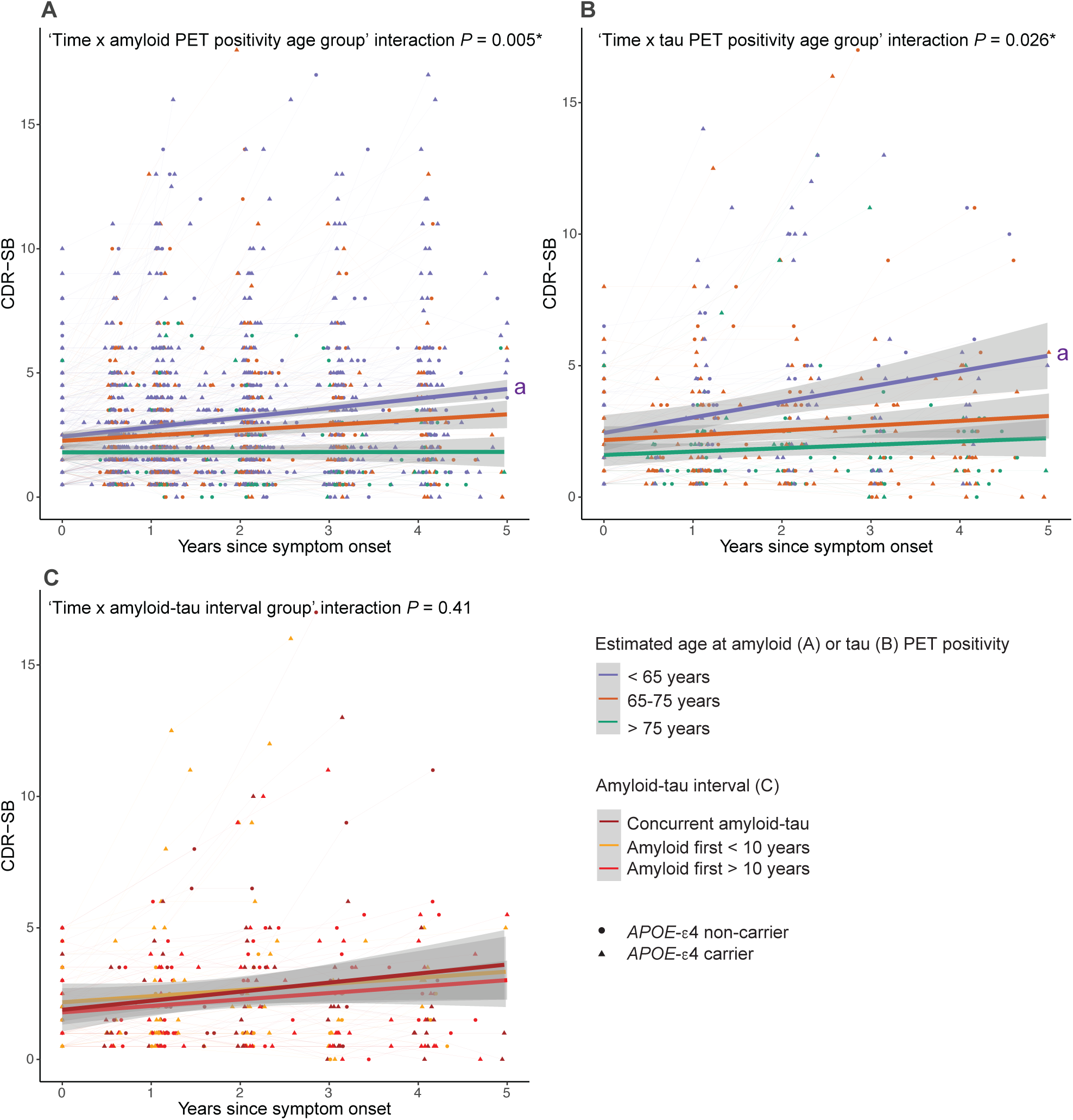
Clinical progression after symptom onset by estimated biomarker positivity age group or amyloid-tau interval group Linear mixed-effects models with random intercepts and slopes were used to model CDR-SB trajectories as a function of years since symptom onset (defined as the first visit with CDR > 0), grouping individuals by estimated age at amyloid PET positivity (A), tau PET positivity (B) or by the amyloid–tau interval (C). Interaction terms between time and biomarker age group or interval group were evaluated to assess differences in the rate of cognitive decline. Pairwise contrasts were conducted to compare group-specific cognitive decline rates. Individual trajectories are shown, where each line connects multiple observations from the same individual. In C, the tau-first group was excluded from the analyses due to the small sample size (n=6). *Statistically significant (*P*<0.05); ^a^ Statistically significant (*P*<0.05) *vs*. ‘>75 years’ group

No significant modifying effects of sex or *APOE*-ε,4 were found in these associations between biomarker positivity age groups or interval groups on the rate of cognitive decline (**eTable 4 in Supplement 1**).

## Discussion

This study investigated the timing and sequence of amyloid and tau PET positivity, and their associations with symptom onset and clinical progression, specifically considering *APOE*-ε4 status and sex differences. Our main findings were: 1) *APOE-*ε,4 carriers and women were younger when they became amyloid and tau PET positive; 2) *APOE-*ε,4 carriers, women, and individuals with older amyloid PET positivity ages had a shorter time from amyloid to tau PET positivity; 3) individuals who became amyloid and tau PET positive at older ages had a shorter time until cognitive symptoms but showed slower rates of cognitive decline after symptom onset; and 4) a shorter time between amyloid and tau PET positivity was associated with a shorter time until symptom onset, but this effect diminished once tau pathology emerged. These results contribute to our understanding of factors influencing heterogeneity in disease progression after amyloid PET positivity, with implications for identifying high-risk individuals for prevention and early therapeutic trials.

Both *APOE-*ε4 carriership and female sex were independently associated with younger amyloid and tau PET positivity ages, consistent with their established AD vulnerability^6–9^. The sex effect was more pronounced among *APOE-*ε4 carriers for amyloid PET positivity, suggesting that female *APOE-*ε4 carriers may be at greatest risk for early development of AD pathology. While interactions between sex and *APOE*-ε4 have been reported for tau vulnerability^10,13–15^, we did not observe a statistically significant effect in our sample, potentially due to limited statistical power or the use of a mesial-temporal meta-ROI capturing very early tau accumulation, where sex differences may not yet be apparent. As expected, estimated amyloid and tau PET positivity ages were highly correlated, reflecting the sequential accumulation of AD pathology^1,2^. This association was significantly stronger in *APOE*-ε4 carriers than in non-carriers, suggesting a tighter coupling between amyloid and tau pathology in genetically at-risk individuals.

A key aim of our study was to evaluate time between positivity by amyloid and tau PET (i.e. the amyloid-tau interval) and its implications for clinical progression. We found that most participants followed an amyloid-first trajectory, consistent with the canonical AD pathway^1,2^. Regarding factors affecting the amyloid-tau interval, *APOE-*ε4 carriers and women exhibited a significantly shorter interval, indicating faster progression to tau pathology after amyloid positivity. Combined with their earlier biomarker positivity ages, these findings reinforce their greater vulnerability to AD pathology. Importantly, older amyloid PET positivity age was associated with a shorter amyloid-tau interval, which may reflect higher prevalence of co-pathologies with increasing age^28–32^ or reduced brain resistance at older ages^33^ that accelerate tau spread after amyloid accumulation.

The observation that older biomarker positivity age was associated with accelerated progression in early disease stages was consistent across both biological and clinical domains. Individuals who became amyloid and tau PET positive at an older age not only exhibited a shorter amyloid-tau interval but also experienced earlier symptom onset. However, following onset of symptoms, they showed slower rates of cognitive decline compared to those with younger biomarker positivity ages. This finding highlights a potential disconnect between vulnerability to symptom onset and subsequent rate of decline. One possible explanation is that younger individuals may show greater cognitive reserve or resilience mechanisms that delay the emergence of clinical symptoms despite pathology, but once a certain pathology threshold is crossed, decline accelerates more rapidly. Notably, our finding of a steeper cognitive decline in individuals with earlier biomarker positivity ages is consistent with previous reports of faster progression in early-onset AD (EOAD) compared to late-onset AD (LOAD)^34–36^.

Beyond the effects of the age at biomarker positivity, a shorter time until tau PET positivity following amyloid PET positivity was associated with an earlier onset of symptoms. Conversely, the amyloid-tau interval duration did not influence symptom onset after tau PET positivity, or rates of cognitive decline. These findings support tau pathology as a primary driver of symptom onset in AD^17–20^ and suggest that once tau pathology emerges, clinical decline proceeds similarly, regardless of when amyloid PET positivity occurred.

Of interest, a longer time until symptom onset following biomarker positivity was observed in women compared to men. However, adjusting for women’s earlier biomarker positivity age largely explained this apparent female advantage, suggesting they experience earlier pathological changes but maintain cognitive resilience longer^37,38^. These findings highlight a sex-dependent vulnerability and the importance of considering both sex and biomarker positivity age when assessing disease progression. In contrast, no differences by sex or *APOE*-ε,4 status were found in the effect of biomarker positivity age or the amyloid-tau interval on rates of cognitive decline after symptom onset, although these analyses were limited by smaller sample sizes and should be further addressed in future studies.

Overall, our findings contribute to a better understanding of the heterogeneity in the amyloid–tau relationship and how it shapes disease progression^4,39–41^. The key contribution of this study is to demonstrate that the age at amyloid and tau PET positivity varies by sex and *APOE*-ε4, and that time between amyloid and tau PET positivity varies considerably and is strongly influenced by age. Further, once tau pathology emerges, it becomes the primary driver of clinical decline, irrespective of when amyloid started to accumulate.

Importantly, our findings are consistent and complement those of recent work^42^ showing that at symptomatic stages, age was negatively associated with prevalence of tau positivity, possibly reflecting age-related heterogeneity in AD pathophysiology, where younger individuals follow a more tau-driven and aggressive trajectory, while older individuals may exhibit symptoms with less tau involvement or alternative non-AD processes. In line with our results, *APOE*-ε4 carriership and female sex were associated to younger ages at amyloid and tau positivity. Another recent study^43^ evaluated factors influencing estimated age at tau positivity. Consistent with our results, they reported that higher amyloid burden and *APOE-ε4* were associated with younger estimated age at tau positivity. They also found that older age at tau PET positivity associated with shorter time from tau PET positivity to dementia. However, unlike our results, they observed an association between low education and literacy and earlier tau positivity and did not report sex differences. Several methodological differences may account for these discrepancies, including variations in the biological clock modeling approach, our study’s use of an earlier tau PET meta-ROI to define the tau timeline, and differing clinical outcome definitions. Specifically, our study focused on the symptom onset rather than dementia stage, as it is a more pertinent milestone for preventive and early-intervention AD trials.

This study has several limitations. First, estimated biomarker positivity ages are dependent on threshold definitions and PET tracers. Second, smaller sample sizes for analyses relative to years from tau PET positivity and the amyloid-tau interval in cognitively impaired individuals limited statistical power, particularly for interaction terms. Third, the low frequency of individuals reaching advanced disease stages limits generalizability to later AD stages, though it enhances translational relevance for prevention and early intervention trials. Finally, heterogeneity in timing of biomarker positivity and subsequent disease progression, influenced by factors such as comorbidities, co-pathologies, lifestyle factors, or social determinants of health, warrant further investigation.

## Conclusions

In summary, this study evaluated how estimated ages at amyloid and tau PET positivity and their temporal sequence contribute to symptom onset and clinical progression, and differ by *APOE-*ε4 status and sex. We found that older age at biomarker positivity and a shorter interval between amyloid and tau PET positivity were associated with earlier symptom onset. Older age at biomarker positivity was also associated with slower subsequent cognitive decline. These findings are crucial for predicting “when” an amyloid-positive individual is likely to develop tau pathology and cognitive impairment, optimizing participant selection for AD clinical trials targeting early disease stages. The influence of *APOE-*ε4 and sex on these temporal dynamics underscores their importance in modeling disease trajectories and designing preventive and therapeutic interventions.

## Data Availability

Data from this study and the study methodology report may be accessed from the ADNI Laboratory of NeuroImaging (LONI) database: adni.loni.usc.edu. The annotated code used for study analyses is provided in full.

https://adni.loni.usc.edu/

## Author Contributions

Concept and design: Milà-Alomà, Schindler, Tosun

Acquisition, analysis, or interpretation of data: All authors

Drafting of the manuscript: Milà-Alomà, Tosun

Critical review of the manuscript for important intellectual content: All authors

Statistical analysis: Milà-Alomà, Hausle, Tosun

Obtained funding: Milà-Alomà, Tosun

## Conflict of Interest Disclosures

Dr. Schindler has served on scientific advisory boards on biomarker testing and education for Eisai and Novo Nordisk and has received speaking fees for presentations on biomarker testing from Eisai, Eli Lilly, and Novo Nordisk. Dr. Tosun reported grants from National Institutes of Health during the conduct of the study.

## Funding/Support

Dr. Milà-Alomà receives funding from Alzheimer’s Association Research Fellowship grant program (AARF-23-1141384). This work was supported by National Institutes of Health (NIH) grants (U19 AG024904, R01 AG091657, U01 AG068057, and U24 AG074855) to Dr. Tosun.

## Acknowledgements

We would like to acknowledge ADNI for ADNI participant data analyzed in this study. Data collection and sharing for the Alzheimer’s Disease Neuroimaging Initiative (ADNI) is funded by the National Institute on Aging (National Institutes of Health Grant U19 AG024904). The grantee organization is the Northern California Institute for Research and Education. In the past, ADNI has also received funding from the National Institute of Biomedical Imaging and Bioengineering, the Canadian Institutes of Health Research, and private sector contributions through the Foundation for the National Institutes of Health (FNIH) including generous contributions from the following: AbbVie, Alzheimer’s Association; Alzheimer’s Drug Discovery Foundation; Araclon Biotech; BioClinica, Inc.; Biogen; Bristol-Myers Squibb Company; CereSpir, Inc.; Cogstate; Eisai Inc.; Elan Pharmaceuticals, Inc.; Eli Lilly and Company; EuroImmun; F. Hoffmann-La Roche Ltd and its affiliated company Genentech, Inc.; Fujirebio; GE Healthcare; IXICO Ltd.; Janssen Alzheimer Immunotherapy Research & Development, LLC.; Johnson & Johnson Pharmaceutical Research & Development LLC.; Lumosity; Lundbeck; Merck & Co., Inc.; Meso Scale Diagnostics, LLC.; NeuroRx Research; Neurotrack Technologies; Novartis Pharmaceuticals Corporation; Pfizer Inc.; Piramal Imaging; Servier; Takeda Pharmaceutical Company Limited; and Transition Therapeutics.

## Notes

### Author Declarations

Data from this study and the study methodology report may be accessed from the ADNI Laboratory of NeuroImaging (LONI) database: adni.loni.usc.edu.

