## Supplement 1 for "Temporal sequence of amyloid and tau PET positivity: *APOE*-ε4 and sex effects, and implications for Alzheimer’s disease progression"

### Supplementary Materials

#### eMethods

Estimates for the age at amyloid and tau PET positivity

To model the biological clock for amyloid and tau based on longitudinal PET observations, we first determined individual rates of change for both amyloid and tau using linear mixed effect (LME) models. These rates were modeled as a function of the pathology burden at the midpoint of the follow-up period (midpoint SUVR). For amyloid PET, separate LME models were run for $^{18}$F-florbetapir (FBP) and for $^{18}$F-florbetaben (FBB) data.

Generalized Additive Models (GAMs) were then used to model amyloid and tau rates of change as a function of midpoint SUVR (Supplementary Figures 1A, 1D, and 1G). To ensure optimal model fit and minimize the influence of high variable data points, we assessed the variance of the GAM fitted values. A cutoff at the 90th percentile of the variance distribution was established to identify and exclude high-variance values (Supplementary Figures 1B, 1E, and 1H). This exclusion resulted in specific SUVR intervals for clock modeling: 0.62–1.11 SUVR for FBP amyloid, 0.63-1.19 for FBB amyloid, and 0.98–2.04 SUVR for FTP tau (Supplementary Figures 1C, 1F, and 1I). Within these refined SUVR intervals, the time corresponding to each 0.0001 SUVR unit increases was calculated by integrating the inverse of the modeled rates of change.

In this study, amyloid time and tau time were conceptualized as the estimated time since PET positivity. For amyloid, a global cortical amyloid PET SUVR of 0.78 for FBP and of 0.74 SUVR for FBB was defined as time 0. For tau, a mesial-temporal meta-ROI tau PET SUVR of 1.41 was set as time 0. Therefore, amyloid or tau time at any given SUVR value was calculated by subtracting the cumulative estimated time at the positivity threshold from the cumulative estimated time at that SUVR value.

To validate our amyloid and tau PET clock models, participants who converted from a negative to a positive PET status during their participation in ADNI study were identified (PET converters; n=81 for FBP amyloid, n=14 for FBB amyloid, and n= 35 for FTP tau). Their actual age at conversion was calculated by averaging their ages at the last negative scan and at the first positive scan. We then tested the associations between the estimated time from the clock models and actual time since conversion; and estimated time intervals between scans (amyloid or tau time at a scan minus amyloid or tau time at first scan) and the actual time intervals between scans.

The estimated amyloid time strongly correlated with actual time since conversion in amyloid PET converters (Spearman's $\rho = 0.91$, $P<0.0001$ for FBP amyloid PET and Spearman's $\rho = 0.74$, $P<0.0001$; Supplementary Figures 2A and 2D). Similarly, the correlation between the estimated tau time and the actual time since conversion in tau PET converters was strong (Spearman's $\rho = 0.76$, $P<0.0001$; Supplementary Figure 2G). The correlations between estimated and actual time intervals were also significant (FBB amyloid time: Spearman's $\rho = 0.87$, $P<0.0001$, Supplementary Figure 2B; FBP amyloid time: Spearman's $\rho = 0.80$, $P<0.0001$, Supplementary Figure 2E; tau time: Spearman's $\rho = 0.58$, $P<0.0001$, Supplementary Figure 2H).

Age at amyloid PET positivity was estimated for all individuals with at least one amyloid positive scan (n=792) by subtracting the corresponding amyloid time from their age at the positive scan. For participants with multiple positive scans, estimated ages were averaged. In amyloid PET converters, the correlation between the estimated age at amyloid PET positivity and actual age at conversion was high (FBB amyloid: Spearman's $\rho$ = 0.76, $P<0.0001$; FBP amyloid: Spearman's $\rho$ = 0.96, $P<0.0001$; Supplementary Figures 2C and 2F). The mean absolute error (MAE) for estimated amyloid age was 2.42 years for FBB and 1.63 years for FBP.

Age at tau PET positivity was estimated for all tau PET scans in participants with at least one positive tau PET scan (n=212). The correlation between estimated age at tau PET positivity and actual age at conversion in tau PET converters was high (Spearman's $\rho$ = 0.96, $P<0.0001$; Supplementary Figure 2I), with an MAE of 2.3 years.

Additionally, the test-retest reliability of the estimated ages at amyloid and tau PET positivity was assessed using the median slope of the estimated ages at positivity across SUVR from different positive scans in individuals with multiple positive scans. Median slopes were -0.23 (IQR:-1.30 to 5.03) for FBB amyloid, -0.29 (IQR: -0.52 to 0.09) for FBB, and -0.52 (IQR: -1.16 to -0.17) for tau PET positivity age.

For subsequent analyses, years from amyloid PET positivity and tau PET positivity were calculated by subtracting the estimated age at PET positivity from the age at the outcome measure.

**eTable 1. Participant characteristics by estimated age at biomarker positivity**

| | Participant characteristics by estimated age at biomarker positivity | | | | | | | |
|---|---|---|---|---|---|---|---|---|
| | Estimated age at amyloid PET positivity | | | | Estimated age at tau PET positivity | | | |
| | **< 65** (n=467; 59.0%) | **65 - 75** (n=225; 28.4%) | **> 75** (n= 100; 12.6%) | ***P*-value** | **< 65** (n=68; 32.1%) | **65 – 75** (n=81; 38.2%) | **> 75** (n= 63; 29.7%) | ***P*-value** |
| Women (n, %) | 253 (54.2) | 108 (48.0) | 38 (38.0) | 0.009* | 41 (60.3) | 36 (44.4) | 26 (41.3) | 0.059 |
| *APOE*-ε4 carriers (n, %) | 344 (43.4) | 113 (50.2) | 36 (36.0) | <0.0001* | 55 (80.9) | 47 (58.0) | 25 (39.7) | <0.0001* |
| Educational attainment (High school or less / College / Postgraduate), n (%) | 73 (15.6) / 213 (45.6) / 181 (38.8) | 33 (14.7) / 86 (38.2) / 106 (47.1) | 14 (14.0) / 51 (51.0) / 35 (35.0) | 0.15 | 6 (8.8) / 40 (58.8) / 22 (32.4) | 10 (12.3) / 34 (42.0) / 37 (45.7) | 4 (6.3) / 29 (46.0) / 30 (47.6) | 0.21 |
| Race (Black/White/Other), n (%) | 26 (5.6) / 424 (90.8) / 17 (3.6) | 8 (3.6) / 211 (93.8) / 6 (2.7) | 5 (5.0) / 93 (93.0) / 2 (2.0) | 0.67 | 6 (8.8) / 59 (86.8) / 3 (4.4) | 0 (0.0) / 75 (92.6) / 6 (7.4) | 2 (3.2) / 55 (87.3) / 6 (9.5) | 0.06 |
| Estimated age at amyloid PET positivity, years | 55.8 ± 6.26 | 69.6 ± 3.08 | 79.7 ± 3.90 | <0.0001* | 53.1 ± 8.36 | 60.4 ± 6.50 | 71.6 ± 6.46 | <0.0001* |
| Estimated age at tau PET positivity, years | 65.1 ± 7.68 | 75.7 ± 7.39 | 80.4 ± 7.22 | <0.0001* | 58.8 ± 4.96 | 70.1 ± 2.94 | 81.5 ± 4.66 | <0.0001* |
| CDR-SB | 2.05 ± 2.24 | 1.47 ± 1.90 | 0.95 ± 1.63 | <0.0001* | 1.88 ± 1.70 | 2.02 ± 2.77 | 1.51 ± 2.02 | 0.38 |
| CDR (0/0.5/>0.5) at baseline, n (%) | 108 (23.1) / 295 (63.2) / 64 (13.7) | 82 (36.4) / 121 (53.8) / 22 (9.8) | 58 (58.0) / 37 (37.0) / 5 (5.0) | <0.0001* | 15 (22.1) / 47 (69.1) / 6 (8.8) | 31 (38.3) / 36 (44.4) / 14 (17.3) | 26 (41.3) / 30 (47.6) / 7 (11.1) | 0.023* |
| CDR (0/0.5/>0.5) at last visit, n (%) | 86 (18.4) / 257 (55.0) / 124 (26.6) | 84 (37.3) / 97 (43.1) / 42 (18.6) | 48 (48.0) / 43 (43.0) / 9 (9.0) | <0.0001* | 14 (20.6) / 38 (55.9) / 16 (23.5) | 20 (24.7) / 44 (54.3) / 17 (21.0) | 23 (36.5) / 32 (50.8) / 8 (12.7) | 0.18 |
| Number of observations | 4.8 ± 2.9 | 5.8 ± 3.4 | 6.9 ± 4.04 | 0.28 | 3.8 ± 2.8 | 5.4 ± 3.7 | 7.7 ± 4.4 | 0.31 |

Participant characteristics by age at biomarker positivity. CDR data correspond to the first or last PET acquisition visit, as indicated. Data is shown as mean ± standard deviation or n (%). Differences between groups were tested using t-tests for continuous variables and Chi-squared test for categorical variables. *Statistically significant (*P*<0.05)

**eTable 2. Participant characteristics by amyloid-tau interval group**

| | Participant characteristics by amyloid-tau interval group | | | | |
|---|---|---|---|---|---|
| | **Tau-first** (n=6; 3.9%) | **Concurrent amyloid-tau** (n=34; 22.4%) | **Amyloid-first** (n=112; 73.7%) | | ***P*-value** |
| | | | **< 10 years (n=47; 42.0%)** | **> 10 years (n= 65; 58.0%)** | |
| Women (n, %) | 5 (83.3) | 16 (47.1) | 24 (51.1) | 29 (44.6) | 0.33 |
| *APOE*-ε4 carriers (n, %) | 3 (50.0) | 23 (67.6) | 39 (83.0) | 39 (60.0) | 0.05 |
| Educational attainment (High school or less / College / Postgraduate), n (%) | 0 (0.0) / 1 (16.7) / 5 (83.3) | 3 (8.8) / 16 (47.1) / 15 (44.1) | 3 (6.4) / 28 (59.6) / 16 (34.0) | 11 (16.9) / 29 (44.6) / 25 (38.5) | 0.13 |
| Race (Black/White/Other), n (%) | 0 (0.0) / 6 (100.0) / 0 (0.0) | 2 (5.9) / 31 (91.2) / 1 (2.9) | 4 (8.5) / 40 (85.1) / 3 (6.4) | 0 (0.0) / 59 (90.8) / 6 (9.2) | 0.27 |
| CDR-SB | 0.83 ± 0.82 | 1.31 ± 1.44 | 1.37 ± 1.42 | 1.03 ± 1.60 | 0.53 |
| CDR (0/0.5/>0.5) at baseline, n (%) | 2 (33.3) / 4 (66.7) / 0 (0.0) | 11 (32.3) / 21 (61.8) / 2 (5.8) | 13 (27.7) / 32 (68.0) / 2 (4.3) | 29 (44.6) / 33 (50.8) / 3 (4.6) | 0.52 |
| CDR (0/0.5/>0.5) at last visit, n (%) | 2 (33.3) / 4 (66.7) / 0 (0.0) | 6 (17.6) / 20 (58.8) / 8 (23.5) | 10 (21.3) / 29 (61.7) / 8 (17.0) | 15 (23.1) / 39 (60.0) / 11 (16.9) | 0.85 |
| Estimated age at amyloid PET positivity, years | 71.7 ± 7.82 | 66.7 ± 9.36 | 60.2 ± 9.52 | 57.3 ± 9.35 | <0.0001* |
| Estimated age at tau PET positivity, years | 63.7 ± 8.85 | 67.4 ± 9.85 | 67.4 ± 9.34 | 71.6 ± 9.10 | 0.027* |
| Number of observations | 7.1 ± 4.4 | 6.3 ± 4.1 | 4.8 ± 3.6 | 5.2 ± 3.2 | 0.72 |

Participant characteristics by amyloid-tau interval group. CDR data correspond to the first or last PET acquisition visit, as indicated. Data is shown as mean ± standard deviation or n (%). Differences between groups were tested using t-tests for continuous variables and Chi-squared test for categorical variables. *Statistically significant ($P$<0.05)

**eTable 3. Characteristics of cognitively impaired participants at first CDR > 0 visit**

| | Cohort with estimated age at amyloid PET positivity (n= 482) | Cohort with estimated age at tau PET positivity (n=131) | Cohort with estimated PET positivity age for both amyloid and tau (n=96) |
|---|---|---|---|
| Age, years | 74.7 ± 7.2 | 74.9 ± 7.1 | 74.63 ± 6.9 |
| Women, n (%) | 217 (45.0) | 56 (42.7) | 43 (44.8) |
| *APOE*-ε4 carriers, n (%) | 315 (65.4) | 73 (55.7) | 60 (62.5) |
| Educational attainment (High school or less / College / Postgraduate), n (%) | 89 (18.5) / 210 (43.6) / 183 (38.0) | 13 (9.9) / 61 (46.6) / 57 (43.5) | 11 (11.5) / 45 (46.9) / 40 (41.7) |
| Race (Black/White/Other), n (%) | 16 (3.3) / 456 (94.6) / 10 (2.1) | 3 (2.3) / 120 (91.6) / 8 (6.1) | 3 (3.1) / 87 (90.6) / 6 (6.2) |
| CDR 0/0.5/>0.5, n (%) | 0 (0.0) / 428 (88.8) / 54 (11.2) | 0 (0.0) / 123 (90.4) / 13 (9.6) | 0 (0.0) / 66 (68.8) / 30 (31.2) |
| CDR at last visit 0/0.5/>0.5, n (%) | 0 (0.0) / 293 (60.8) / 189 (39.2) | 0 (0.0) / 118 (90.1) / 13 (9.9) | 0 (0.0) / 91 (94.8) / 5 (5.2) |
| CDR-SB | 2.17 ± 1.59 | 1.98 ± 1.54 | 1.79 ± 1.28 |
| CDR-SB at last visit | 4.18 ± 3.3 | 3.90 ± 3.6 | 3.6 ± 3.4 |
| Amyloid PET, Centiloid | 79.1 ± 31.0 | 70.5 ± 46.8 | 75.0 ± 31.2 |
| Amyloid PET-positive[a], n (%) | 390 (94.9) | 96 (81.4) | 83 (93.3) |
| Mesial-temporal tau PET, SUVR | 1.71 ± 0.44 | 1.71 ± 0.26 | 1.73 ± 0.28 |
| Mesial-temporal tau PET-positive[b], n (%) | 70 (73.7) | 82 (94.3) | 57 (93.4) |
| Estimated age at amyloid PET positivity, years | 61.2 ± 9.8 | 60.7 ± 9.6 | 60.7 ± 9.6 |
| Amy age group < 65 / 65-75 / > 75, n (%) | 317 (65.8) / 118 (24.5) / 47 (9.8) | 67 (69.8) / 20 (20.8) / 9 (9.4) | 67 (69.8) / 20 (20.8) / 9 (9.4) |
| Estimated age at tau PET positivity, years | 68.32 ± 8.99 | 68.99 ± 9.24 | 68.32 ± 8.99 |
| Tau age group < 65 / 65-75 / > 75, n (%) | 36 (35.6) / 37 (38.5) / 23 (24.0) | 45 (34.4) / 50 (38.2) / 36 (27.5) | 36 (37.5) / 37 (38.5) / 23 (24.0) |
| Amyloid-tau interval | - | - | 7.6 ± 6.9 |
| Amyloid-tau interval group (Tau first / Concurrent amyloid-tau / Amyloid first < 10 years / Amyloid first > 10 years) | - | - | 5 (5.3) / 22 (22.9) / 32 (33.3) / 37 (38.5) |

Data are presented as mean ± standard deviation or n (%). Participants were classified as cognitively impaired if they had two consecutive visits with a CDR > 0 and a CDR > 0 at their last visit. Data from their first visit with a CDR > 0 is reported unless otherwise specified. 411 and 95 participants with estimated amyloid PET positivity age had an amyloid PET and a tau PET within a year of their first CDR 0 visit, respectively. 118 and 87 participants with estimated tau PET positivity age had an amyloid PET and a tau PET within a year of their first CDR 0 visit, respectively. 96 participants had estimates for both amyloid and tau PET positivity age and overlapped between the two cohorts. Of those, 89 and 61 had an amyloid PET and a tau PET within a year of their first CDR 0 visit, respectively. [a]Amyloid PET positivity threshold: > 0.78 SUVR for FBP scans and > 0.74 SUVR for FBB scans; [b]Mesial-temporal tau PET positivity threshold: > 1.41 SUVR.

#### eTable 4. Rate of decline after symptom onset by biomarker positivity age group or amyloid-interval group

| | Rate of change in CDR-SB (CDR-SB/year) | 'Time per group' interaction | | 'Time per group per sex' interaction | | 'Time per group per *APOE*-ε4' interaction | |
|---|---|---|---|---|---|---|---|
| | β [95% CI] | F (df1, df2) | *P*-value | F (df1, df2) | P-value | F (df1, df2) | *P*-value |
| **Amyloid age group** | | 5.40 (2, 349 | 0.005* | 2.63 (2, 348) | 0.07 | 1.19 (2, 355) | 0.31 |
| < 65 | 0.96 [0.84 - 1.09] | | | | | | |
| 65 - 75 | 0.78 [0.57 - 0.98] | | | | | | |
| >75 | 0.41 [0.08 - 0.73][a] | | | | | | |
| **Tau age group** | | 3.77 (2, 102) | 0.026* | 0.29 (2, 97) | 0.75 | 0.82 (2, 99) | 0.44 |
| < 65 | 1.07 0.74 -1.39] | | | | | | |
| 65 - 75 | 0.72 [0.41- 1.02] | | | | | | |
| >75 | 0.39 [0.03 - 0.76][a] | | | | | | |
| **Amyloid tau-interval group** | | 0.98 (3, 76) | 0.41 | 0.71 (3, 70) | 0.55 | 0.98 (3, 73) | 0.41 |
| Concurrent amyloid-tau | 0.83 [0.41 - 1.26] | | | | | | |
| Amyloid first < 10 years | 0.81 [0.44 - 1.18] | | | | | | |
| Amyloid first > 10 years | 0.47 [0.14 - 0.81] | | | | | | |

Change in CDR-SB per year [95% CI] in participants after symptom onset (first visit with a CDR > 0). Participants were classified by age at biomarker positivity group or by amyloid-tau interval group. Linear mixed effect (LME) models with random slopes and intercepts were used to assess the effect of biomarker positivity age or amyloid-tau interval on the rates of cognitive decline over five years following symptom onset Interaction terms between time since symptom onset and biomarker age group or interval group were evaluated. Next, pairwise contrasts were conducted to compare group-specific cognitive decline rates. Separate models included interactions between time, biomarker positivity age or amyloid-tau interval group and sex or *APOE*-ε4 status. In the amyloid-tau interval analyses, the tau-first group was excluded due to the small sample size (n=6).
*Statistically significant ($P$<0.05)
[a] $P$<0.05 *vs.* < 65 years

#### eFigure 1. Model fits to estimate amyloid and tau PET clocks

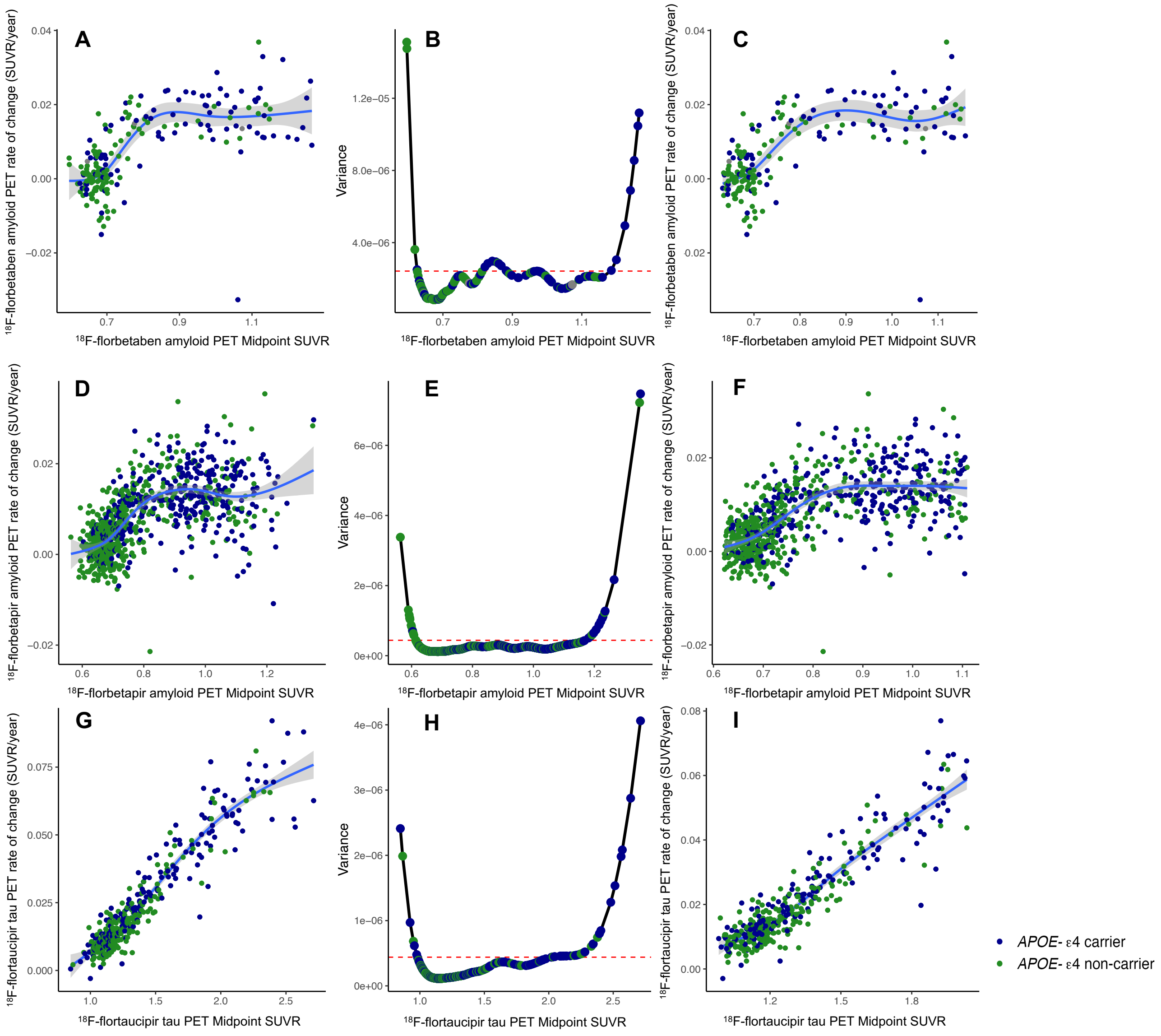

Generalized Additive Models (GAM) with cubic spline were applied to evaluate the relationship between amyloid or tau burden halfway the follow-up period (midpoint SUVR) and the rate of change in amyloid or tau PET. For amyloid PET, $^{18}$F-florbetaben (n=204) and $^{18}$F-florbetapir (n=784) data were modelled separately. A, D and G show the GAM model fit including the entire midpoint SUVR range for amyloid (A and D) and tau PET (G) data. B, E and H show the variance of the GAM fitted values across midpoint SUVR values for amyloid (B and E) and tau PET (H) data. Horizontal dashed red line depicts the 90th percentile variance cutoff. C, F and I show the GAM model fit after excluding midpoint SUVR values above the 90th percentile variance cutoff, which resulted in an interval of 0.63-1.19 for $^{18}$F-florbetaben amyloid PET, 0.62-1.11 SUVR for $^{18}$F-florbetapir amyloid PET, and 0.98-2.04 SUVR for tau PET. Each point depicts the value of an individual and the solid blue lines indicate the model fit. *APOE*-ε4 carriers are shown in blue and *APOE*-ε4 non-carriers are shown in green.

#### eFigure 2. Validation of the amyloid and tau PET clock estimates

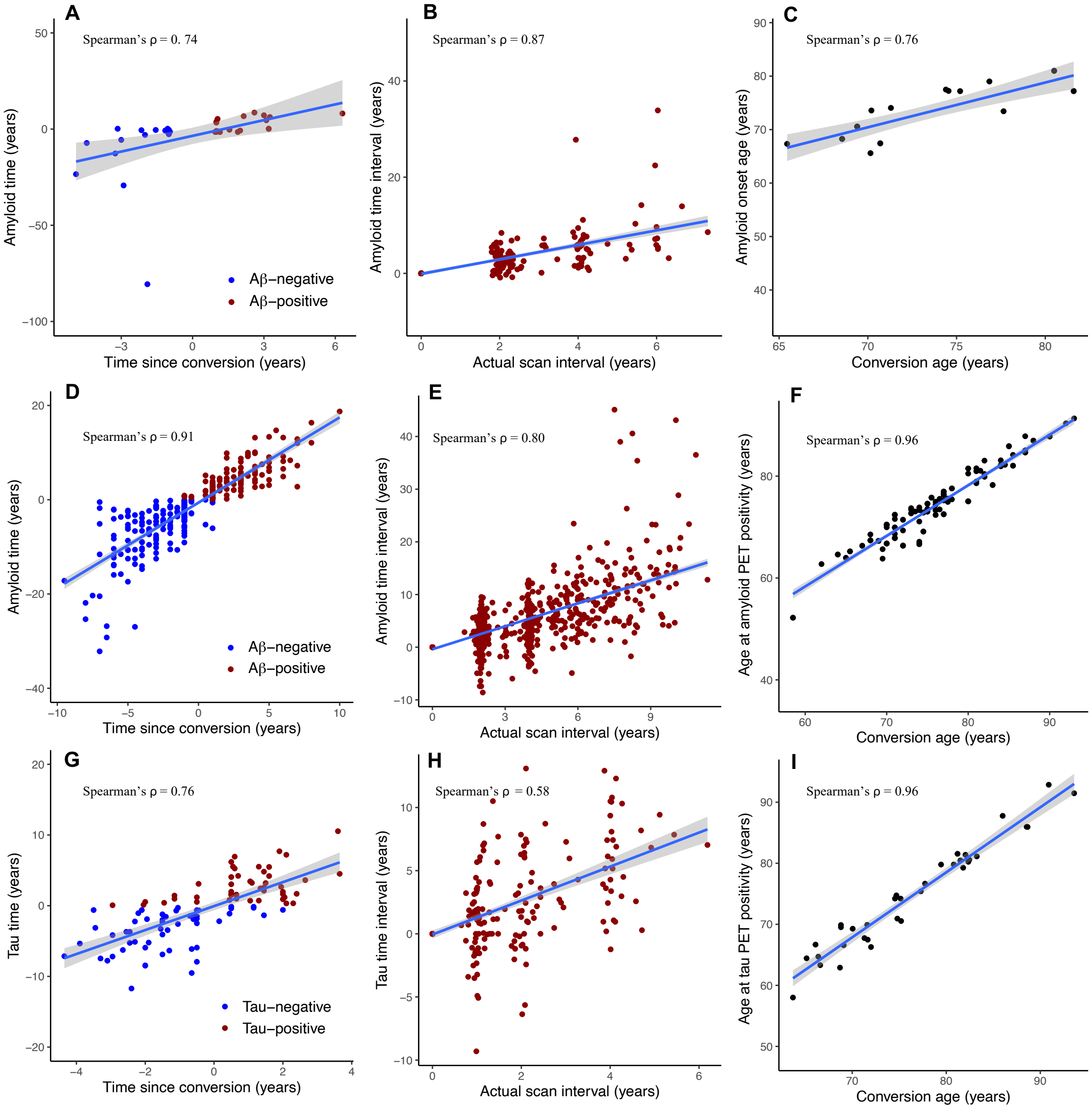

Amyloid and tau time models were validated in the group of individuals that converted from a negative to a positive amyloid or tau PET scan throughout the study duration, respectively. Actual conversion age in PET converters was calculated by averaging the ages at the last negative scan and at the first positive scan. A, D and G show the correlation between the estimated amyloid time (A and D) or tau time (G) and the actual time since conversion. B, E and H show the correlation between amyloid (B and E) or tau (H) time intervals between scans (amyloid or tau time - amyloid or tau time at first scan) and actual time intervals between scans above the positivity threshold in the whole sample. C, F and I show the correlations between the estimated age at amyloid PET positivity (C and F) or tau PET positivity (I) with the actual age at conversion in PET converters. Of note, each point at A, B, D, E, G and H depict a scan and each point at C, F and I depict an individual. Solid lines indicate the linear regression lines.

**eFigure 3. Effect of estimated biomarker positivity age and amyloid-tau interval on time until symptom onset following amyloid or tau PET positivity**

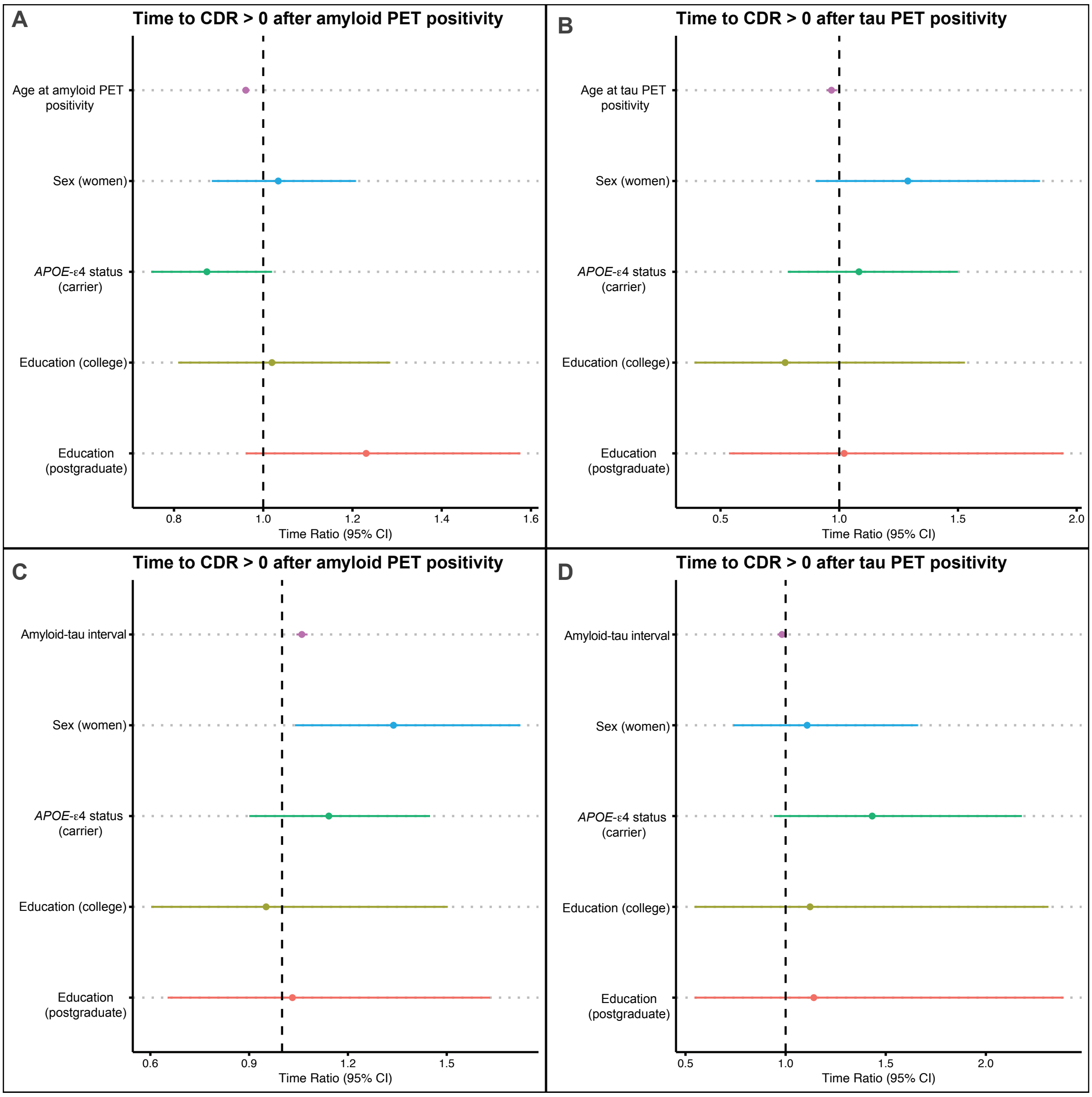

Forest plots displaying time ratios and 95% confidence intervals (CI) from Accelerated Failure Time (AFT) models examining time from amyloid or tau PET positivity to CDR > 0. A time ratio > 1 indicates slower progression (longer time to impairment), whereas a time ratio < 1 indicates faster progression (shorter time to impairment). A and B include the effects of the estimated age at amyloid PET positivity or tau PET positivity on progression rate in the amyloid or tau timeline, respectively. B and C include the effects of the amyloid-tau interval on progression rate in the amyloid or tau timeline, respectively. All models included sex, *APOE*-ε4 status and educational attainment as covariates.
